# Detecting Wolff-Parkinson-White from Lead-I ECG Using Transfer Learning and Wavelets

**DOI:** 10.1101/2024.09.26.24314432

**Authors:** Shreyas Bharadwaj, Sarah Altman, Jessica Wang, Ridwan Alam

## Abstract

Wolff-Parkinson-White (WPW) syndrome is a congenital heart defect that can trigger ventricular fibrillation and sudden cardiac death. Expert inspection of a 12- lead clinical electrocardiogram (ECG) or a Holter record is the standard approach for detecting WPW syndrome. Smartwatches that acquire lead-I ECGs can enable automated detection of WPW episodes in out-of-hospital settings and prevent adverse outcomes. In this work, we explore deep-learning solutions to identify WPW syndromes on lead-I ECG. Scarcity of labeled ECG data for WPW and other cardiac conduction disorders poses a major challenge for training data-driven methods. Moreover, generalizability of such methods to external patient population remains unexplored. To address these challenges, first we implement and compare multiple existing strategies from time-domain augmentations on the lead-I ECG to transfer learning of Imagenet-models for ECG wavelet transformations. Training and holdout validation of these methods are conducted using about 14,000 ECGs from PTB-XL, a publicly available ECG dataset. Moreover, we explore generalization of these methods by external validation on the data from 140 patients from the Tongji Hospital ECG Database. While these methods achieve 88% sensitivity and 99% specificity in identifying a lead-I ECG with evident WPW, and an area under the receiver-operating curve (AUC) of 0.99 on the holdout set from PTB-XL, the sensitivity drops to 58% with an AUC of 0.88 on the external validation. Finally, we propose a novel data augmentation strategy by incorporating labeled data from an umbrella super-class of cardiac conduction disorders, instead of WPW alone, thus naturally reducing the data imbalance for model training. We apply these models as zero-shot transfer learning for discriminating WPW from normal ECG. While this approach achieves similar performance during holdout validation, it also demonstrates strong performance on the external Tongji dataset with sensitivity 0.78 and AUC 0.91. This result shows significant generalizability of the proposed method and highlights the potential of deep-learning solutions in monitoring WPW syndrome with lead-I ECG in outof-hospital general populace settings.

## I. Introduction

ELECTROMECHANICAL coupling of polarizing cellular currents and resultant myocardial contractions in the heart enables adequate blood supply to the body. Conduction disorders (CDs) disrupt the heart’s sinus electromechanical coupling, resulting in aberrant electrical currents and consequent disjointed contraction. Wolff-Parkinson-White (WPW) syndrome is one such CD, affecting 0.1%-0.3% of the general population [1], [2]. WPW is characterized by accessory electrical pathways which allow impulses to bypass the atrioventricular node when conducting from the atria to the ventricles, facilitating life-threatening conduction of rapid atrial fibrillation impulses to the ventricles [3], [4]. Sudden cardiac death arising from such aberrant conduction has a 3-4% lifetime risk for those with WPW, necessitating accurate diagnostic measures to indicate candidates for curative catheter ablation [5]. Symptomatic WPW patients are additionally predisposed to congestive heart failure, as prolonged tachyarrhythmias can cause left ventricular dilation and systolic dysfunction; importantly, prompt identification and treatment of causal tachyarrhythmic agents can lead to symptom amelioration and improvement of systolic function, highlighting the importance of early WPW diagnosis [6]. However, the episodic nature of WPW, coupled with the lack of visible WPW signs during sinus rhythm, hinders early detection and timely intervention [3], [6].

In practice, the diagnosis of WPW relies on capturing an episode in real-time via ECG, a task complicated by the condition’s transient and often asymptomatic behavior (Fig 1). WPW typically manifests on an ECG through short (*<*120 ms) PR intervals, wide (*>*120 ms) QRS complexes, and delta-wave slurring; the absence of these characteristics, however, does not preclude WPW diagnosis [3].

**Fig. 1:**
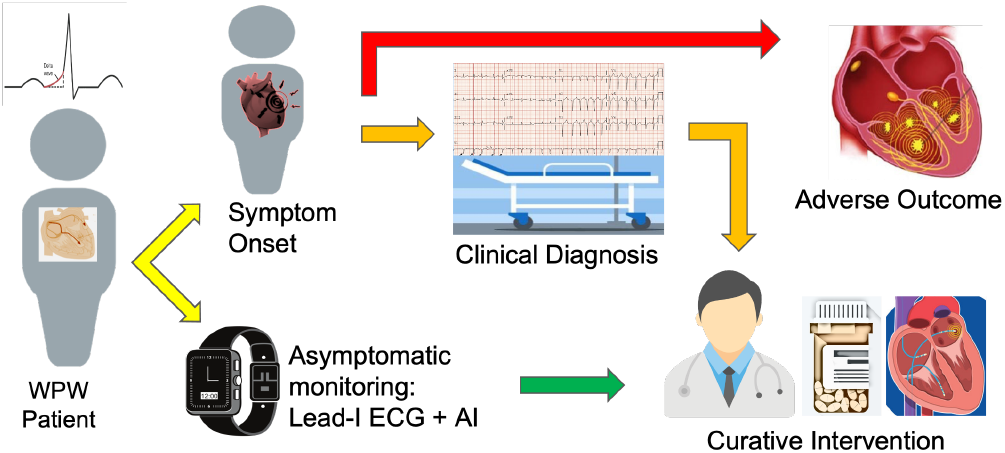
Lead-I ECG monitoring can enable timely WPW detection and improve patient outcomes.

Beyond logistical challenges, the detection of a WPW episode is limited by the accuracy of physician-based ECG assessment in resource-limited settings; without specialized pre-training in ECG analysis, pooled accuracy’s of resident physicians and cardiologists in ECG interpretation approach 55.8% and 74.9%, respectively, underscoring a need for automated arrhythmia detection tools as diagnosis support resources [7].

Traditional algorithms for identifying WPW from 12-lead ECGs have focused on retrospective localization of APs for treatment planning, which limits their utility as proactive diagnostic tools [8], [9]. Machine learning (ML) approaches utilizing ECG edge tracking lack robustness for detecting WPW in cases without delta-wave ECG slurring [10]. Recent efforts utilizing multi-modal deep learning (DL) networks for AP classification in WPW from 12-lead ECGs are limited by poor predictive performance in right anteroseptal and left posteroseptal regions, and are reliant upon large amounts of training data [11], [12]. Notably, the rarity of WPW precludes model training with a high WPW data volume; other conduction disorders (CD), however, experience prevalence up to 20% at baseline in the general populace [18]. CDs such as bundle branch blocks and fasicular blocks present on an ECG with similar clinical findings to WPW, suggesting the utility of training on CD ECGs to address class imbalances in WPW classification tasks.

Given these challenges, the motivation for the present study emerges; there is urgent need for a more reliable, robust automated detection framework for diagnosing WPW pathologies. A substantial limiting factor concerning the clinical utility of current WPW diagnosis algorithms, ML models, and DL models is their reliance upon 12-Lead ECG input. Our objective is to identify whether lead-I alone contains sufficient information for automated WPW detection. Toward the goal, we evaluate two independent methods: firstly, we investigate whether randomly selected existing time-domain augmentation followed by continuous wavelet transformation of the Lead-I signal input can effectively train a deep-learning pipeline to discriminate WPW, provide accurate classifications, and generalize beyond it’s internal dataset. Secondly, we investigate the feasibility of a natural data augmentation for this task by training a model to classify the larger CD superclass, and interrogate its generalizability as a WPW subclass classifier (Fig. 2). In both cases, we aim to explore whether deep neural networks can effectively classify WPW from lead-I ECGs alone; as this claim holds promise for wide-spread WPW disease monitoring in the general populace via ECG-enabled smart devices, transforming the WPW diagnostic landscape to enable timely intervention.

**Fig. 2:**
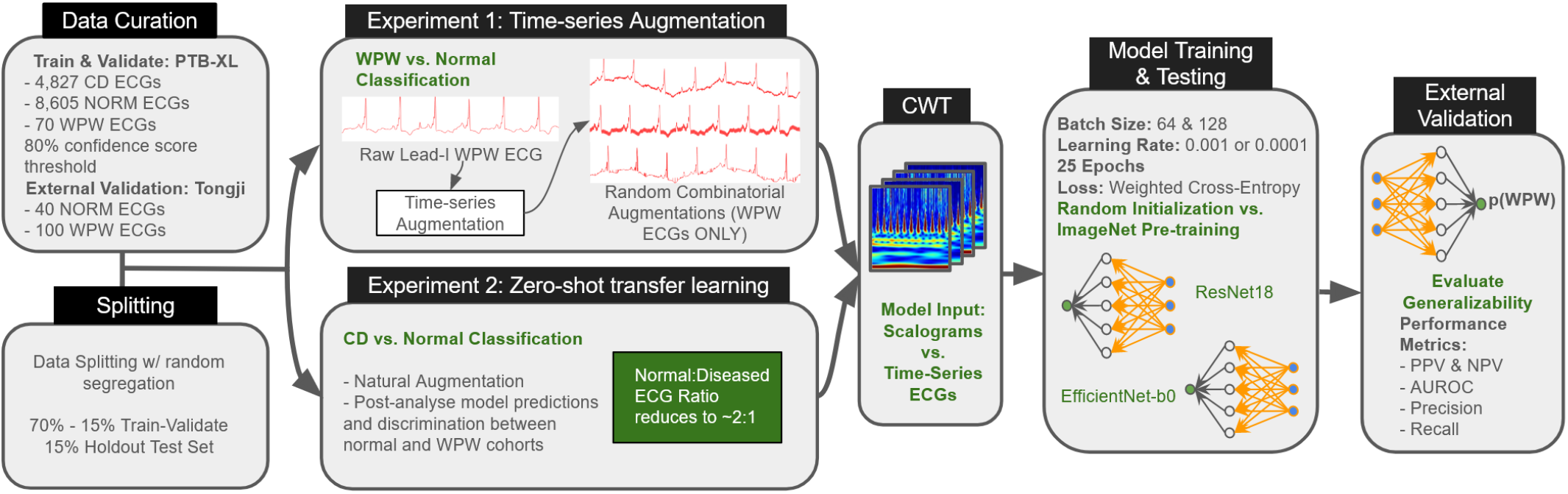
Workflow schematic for model pipeline development. Two WPW vs. Normal classification task experiments were performed in parallel; one with only WPW data in the diseased class, and another using a larger CD superclass for natural augmentation.

**Fig. 3:**
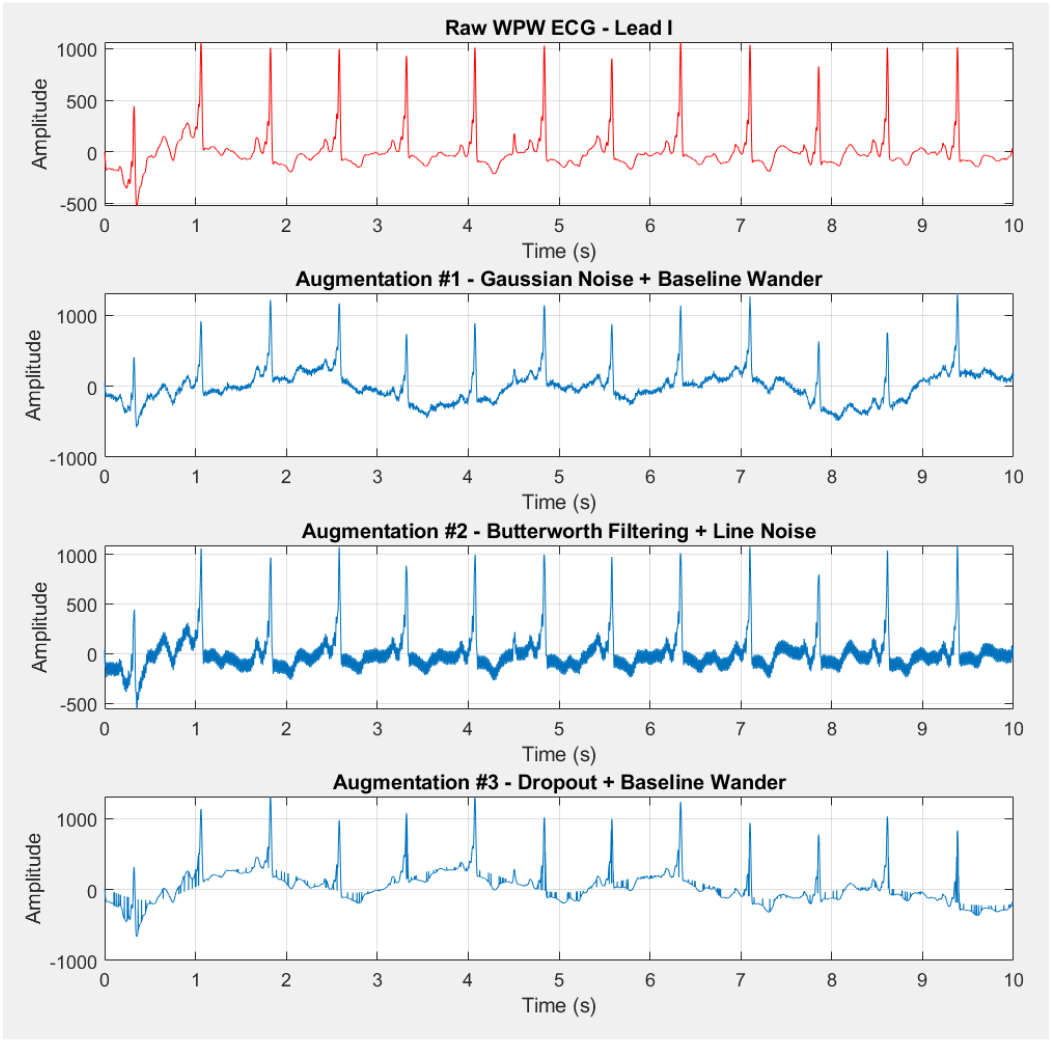
Original lead-I 10-second clinical ECG with evident WPW syndrome and 3 augmented versions of this ECG. The combined augmentations include: 1. Addition of Gaussian noise & baseline wander, 2. Butterworth bandpass filtering & line-noise effect, and 3. Dropout & baseline wander.

## II. Related Works

Traditional clinical algorithms for WPW diagnosis, such as the one by Arruda et al. [8], localize accessory pathways using 12-lead ECG and catheter ablation data, achieving high sensitivity (0.90) and specificity (0.99). However, these methods rely on retrospective data, struggle with multiple pathways or structural heart disease, and are less effective in children. The EASY-WPW algorithm by El Hamriti et al. [9] offers improvements with a sensitivity of 0.92 and specificity of 0.99, including validation in children. Whether such performance would translate to a single-lead setting, as well as to external patient populations, remains unanswered.

Machine learning (ML) and deep learning (DL) have significantly advanced the automated detection of WPW syndrome. Merbouti et al. detail a novel ML-based approach that identifies WPW’s critical Delta wave using several algorithms, including Neural Networks, Näive Bayes, and KNearest Neighbors (KNN) [10]. They achieved an accuracy of 0.9716, but their method relies heavily on precise identification of ECG peaks; this can be problematic for noisy signals, atypical rhythms, or cases with hidden or transient APs. To avoid reliance on explicit feature detection, deep learning (DL) techniques have also been pursued [11], [13]. Zhu et al. [12] used convolutional neural networks on images of timeseries ECG data, achieving high sensitivity and specificity for WPW detection. Strodthoff et al. [14] benchmarked several approaches for multilabel classification using the PTB-XL dataset, including convolutional neural networks with 1D ECG input, LSTMs, wavelet-based neural networks, and ensemble methods. They achieve a holdout AUC of 0.855 in classifying WPW vs. normal ECGs from 12-lead ECG input. This 12-lead performance is not directly comparable to our lead-I ECG-based task, yet we consider this result as a target baseline for our methods. It was noted that although 1D-CNNs performed the best, further investigation is needed to determine utility of wavelet models in particular, due to the sensitivity to preprocessing. Other challenges persist with current deep learning approaches, including reliance on large amounts of time-series training data from all 12 leads. Crucially, these algorithms have also not been validated on external datasets, limiting their generalizability.

The growing availability of ECG-enabled wearable devices presents a potential solution to limitations posed by 12-Lead ECGs [15]. Fiorina et al. demonstrated good detection accuracy using smartwatch-based ECG monitoring [16], and AbuAlrub et al. demonstrated the feasibility of automated atrial fibrillation diagnosis with consumer-grade wearables [17]. These developments suggest a promising future for remote, wearable-enabled lead-I ECG monitoring for WPW detection. Overall, while existing automated techniques have advanced WPW detection, there are significant assumptions and limitations, including reliance on high-quality ECG data, the need for validation across diverse populations, and the challenge of implementing these technologies in resource-limited settings. The potential for wearable devices to provide accessible and reliable ECG monitoring offers a promising avenue for overcoming some of these challenges in WPW diagnosis, and motivates development of robust, generalizable, and proactive monitoring for WPW using only lead-I ECGs.

## III. Methods

### A. Data Description

We use two datasets in this work: the PTB-XL dataset to train and validate our models, and the Tongji Hospital dataset to conduct external validation. While both datasets host 12-lead, 10-second clinical ECGs, we use only Lead-I ECGs for our experiments.

PTB-XL [19] is a large ECG dataset, publicly available on PhysioNet, that contains 21,837 clinical 12-lead ECGs from 18,885 patients; diagnoses include conduction disorders (CD), myocardial infarctions, ischemia, and hypertrophic cardiomyopathy, as well as those without any cardiac disorders. 67.13% of ECG annotations were performed by at least one cardiologist, 31.2% were generated via automatic device interpretation, and 1.67% had no initial annotation; following initial annotation, a random subset of the data was secondarily annotated by an independent cardiologist. For our experiments, we curate a subset of 13,432 ECGs whose labels are validated by at least one cardiologist with confidence scores of at least 80%; included are 4,827 ECGs labeled as positive for one or more CD categories: fascicular blocks, bundle branch blocks, AV blocks, Wolff-Parkinson-White syndromes, or non-specific CDs. Within this CD superclass, only 70 ECGs are labeled as having evidence of WPW syndrome. 8,605 included ECGs are labeled as “normal,” or without any cardiac disorders. The normal patient cohort has mean±std age of 52±22 years (median 53 years), with 55% being female. The CD cohort is 72±42 years old (median 68 years) with 39% female patients. The WPW subset has a distribution of 48±17 years, and 46% of patients are female.

The Tongji Hospital ECG Database [12] is a publicly available resource from the Huazhong University of Science and Technology. It contains labeled standard 12-lead, 10-second ECG recordings from 828 patients. All ECG were annotated by a primary board-certified cardiologist and secondarily validated by senior cardiologists into 21 rhythm subtypes, including conduction disorders and arrhythmias. We utilize 40 ECG samples with “normal” or no-disorder labels and 100 samples with evidence of WPW (type A and B). Normal and WPW patient cohorts have age distributions of 41±13 and 42±14 years, respectively. Demographics related to sex are not available for this dataset.

### B. Experimental Setup

To build our data-driven solution, we split our PTB-XL data with stratification into training-validation-holdout sets using a 70-15-15% split of patients. All ECGs of a particular patient were then assigned to the corresponding training, validation, or holdout set. This was done to avoid data leakage between ECGs of the same patient, and ensure that the validation and holdout sets were as agnostic of training data as possible.

The PTB-XL dataset contains a large imbalance between ECG classes; a more than 1000-to-1 ratio between “normal” ECGs and those with WPW diagnoses exists. Such highly imbalanced data is hard to use for both training and validation of any data-driven model owing to a small number of samples per class and a loss of statistical validity. With the above split, our holdout set contains only 10 WPW samples, introducing challenges with class imbalances.

In this paper, we attempt to counter such imbalance on model performances. Toward that objective, we run two experiments. First, we explore existing time-domain augmentations and continuous wavelet transformation (CWT) of the lead-I signal to increase the size of our WPW training data set four-fold. We then use generated scalograms as input to both randomly initialized and pre-trained models for comparison of WPW classification performance.

Second, we consider all cardiac conduction disorders (CD) as our target disease class (which includes WPW data), thus decreasing the ratio of normal-to-diseased ECGs to 2-to-1. This form of natural augmentation provides a much more reasonable split that can be easily addressed in the model training pipeline with existing strategies such as data augmentation and loss weight-balancing. We then post-analyse this model’s predictions to quantify its performance on discriminating normal vs WPW ECG cohorts. A schematic workflow for the experiments and corresponding decision conditions are presented in Figure 2.

### C. Data Augmentation

Data augmentation can be used to increase training dataset size, and has shown to increase generalizability and reduce bias for ECG-based models [20]. Translations such as the addition of baseline wander and Gaussian noise are commonly used to yield improved model performance; further, random combinations of data augmentations are shown to outperform individual data augmentation operations for ECG classification tasks [21]. Our objective is to utilize existing knowledge in ECG augmentation for addressing data imbalance. Hence, we implement multiple existing augmentation algorithms and randomly pair them to produce 3 augmented lead-I ECGs for our WPW class. This strategy increases our WPW dataset volume four-fold; for the WPW-targeted experiments, the ratio between normal-to-diseased classes reduces to about 400-to-

1. Augmentation was not performed on the normal class or the conduction disturbance superclass, as we have sufficient original samples for those categories.

We implemented five base augmentation operations:

1. Gaussian Noise Addition: Adds white Gaussian noise to ECG with power between 15 and 30 dB.
2. Baseline Wander Injection: Adds a combination of 3 sine functions to the ECG to mimic low-frequency motion and respiration artifacts. Each sine function has a random frequency (range 0.05 Hz to 0.5 Hz), random phase (range 0 to 2*π* rad), and random amplitude (range 5% to 20% of raw input amplitude).
3. Butterworth Filtering: Applies a 5th-order bandpass Butterworth filter with a lowpass of 0.5 Hz and a highpass of 50 Hz to the ECG signal.
4. Line-Noise Effect: Adds a sine function to signal to mimic power line noise, characterized by a frequency of 60 Hz, a random phase between 0 to 2*π* rad, and random scaling (5% - 10% of raw input amplitude).
5. Dropout Elimination: Randomly sets 5% of data points in a segment to zero to mimic data corruption.

Our data augmentation pipeline, implemented in MATLAB, randomly selects three pairs of augmentation functions from the list above, followed by an intra-pair linear combination. Each WPW lead-I ECG is augmented to produce three 10-second signals, and the raw (original) segment is maintained, thereby increasing the WPW training volume to reduce the class imbalance. All lead-I ECGs, regardless of augmentation status, are then converted to scalograms using the continuous wavelet transform.

### D. Continuous Wavelet Transform

Wavelets are pulse-like, time-limited oscillations that offer the ability to capture time and frequency information simultaneously in signal processing. The continuous wavelet transform decomposes a signal into coefficients of wavelets at different scales. Thus, high frequency localized oscillations present in a finite-duration signal are captured along with low frequency oscillations, enabling multi-resolution analysis [23], [24].

For a 1D input, the continuous wavelet transform generates a scalogram, which is a two dimensional log-intensity plot of wavelet coefficient magnitudes (Figure 4). The *x*-axis of the scalogram corresponds to time, and the *y*− axis corresponds to frequency; therefore, the intensity of a given point on the scalogram can be understood as the weightage of a particular wavelet in the signal.

**Fig. 4:**
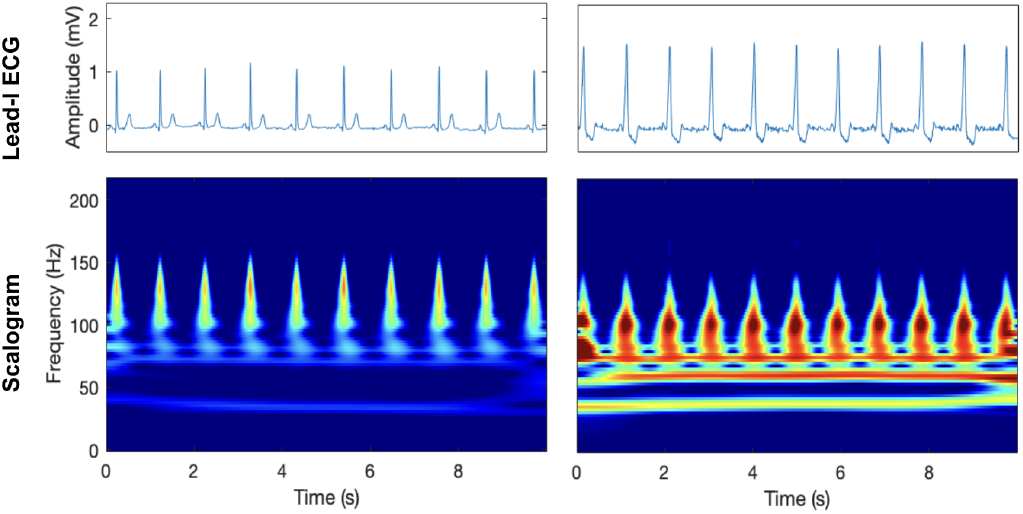
Lead-I ECG and corresponding time-frequency scalograms obtained from wavelet transforms of a patient with no cardiac abnormality (left) and a patient with evident WPW syndromes (right). Notably, the scalograms seem to emphasize the characteristic features of WPW cases, such as short PR intervals and presence of delta waves, potentially adding visual interpretability.

Mathematically, the wavelet transform projects the time-domain signal onto the basis spanned by wavelets at different scales:

where 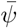 is the complex conjugate of the wavelet, and *a* and *b* describe the scaling and shifting of the wavelet, respectively, that enable multi-scale decomposition of the input signal.

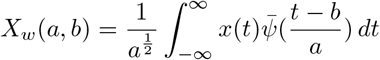

Based on their demonstrated success in wavelet analysis of ECG [23], [24], we applied the continuous wavelet transform using the Generalized Morse wavelet to each ECG sample. This generated a 224*×* 224 *×*3 RGB image to be used as input for the CNN, as shown in Figure 4.

### E. Deep Neural Network Architecture

We implement and compare two state-of-art model archi-tectures, EfficientNet-b0 and ResNet18.

#### EfficientNet-b0

EfficientNet-B0 [25] is a deep convolutional neural network architecture developed for image classification tasks, introduced by Tan and Le in 2019. It employs a novel model scaling method that uniformly scales network depth, width, and resolution using a compound coefficient. EfficientNet-B0 has around 5.3 million parameters and achieves high accuracy with significantly fewer parameters compared to other architectures. On the ImageNet dataset, EfficientNet-B0 attained a top-5 error rate of 2.88%, demonstrating its capability in producing accurate results with lower computational costs.

#### ResNet18

ResNet18 [26] is a deep convolutional neural network architecture designed for image classification and recognition tasks. It comprises 18 layers and around 11 million parameters, utilizing residual learning to address the vanishing gradient problem, which allows for the training of much deeper networks. Each residual block contains two convolutional layers with a shortcut connection that performs identity mapping, ensuring that gradients can flow through the network more effectively. ResNet18 achieved a top-5 error rate of 5.71% on the ImageNet dataset.

To perform binary image classification, we modified each pretrained architectures by replacing the final fully-connected layer to match the number of classes. In EfficientNet, we replaced the third-to-last 2-D global average pooling layer with a dropout layer of probability 0.6. In ResNet18, we replaced the fourth-to-last 2-D global average pooling layer with a dropout layer with probability 0.6. These were included to enhance regularization and discourage overfitting. The modified network architectures are shown in Figure 5.

**Fig. 5:**
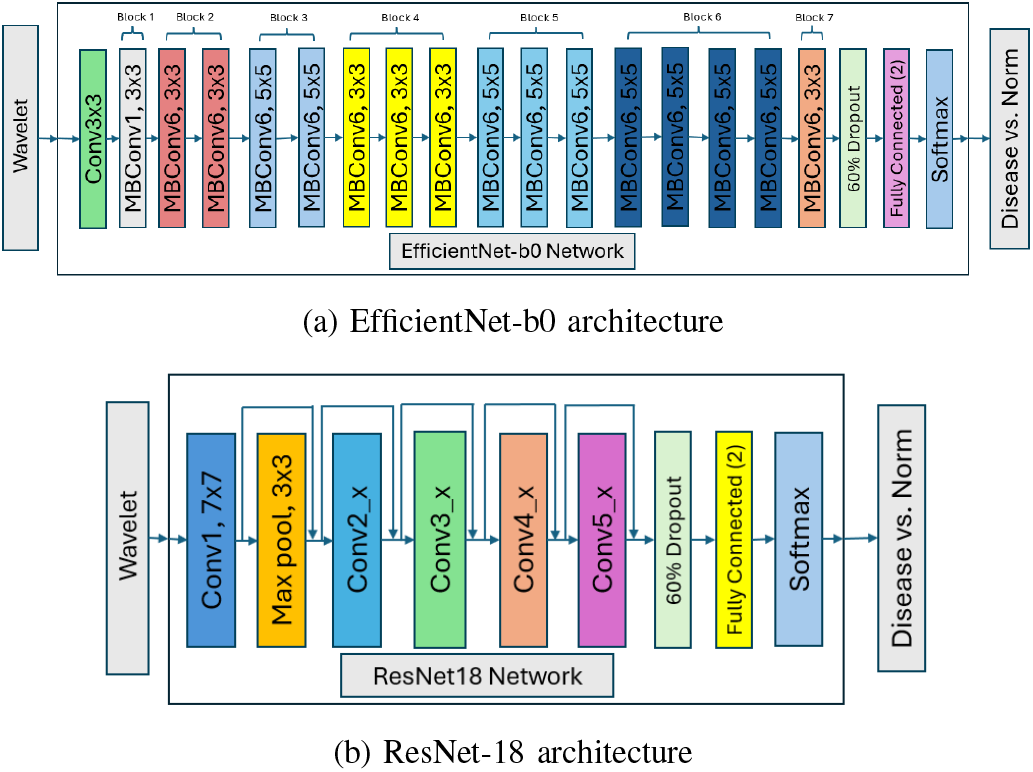
Model architectures: (a) depicts the standard EfficientNet-b0 adopted from [24], consisting of mobile bottleneck inverted convolution (MBConv), dropout, fully-connected, and softmax layers, and (b) refers to the ResNet-18 from [25] that uses convolutional blocks (conv) with residual connections, pooling, dropout, linear, and softmax layers. Both architectures use wavelet scalograms as input and output binary-classification posterior probabilities.

### F. Training

Due to the inherent similarities in features in many image classification tasks, transfer learning has proven extremely effective, especially when training on small datasets [27]. We attempted and compared supervised learning using random initialization with fine-tuning from ImageNet-pretrained networks. For each network architecture, we tested batch sizes of 64 and 128. Learning rates were set to either 0.001 or 0.0001 with a piece-wise step-scheduler that decreased the learning rate by a factor of 0.2 every 5 epochs. Each network was trained for 25 epochs. Weighted cross-entropy loss was used as optimization criteria to account for class imbalances.

### G. Evaluation Metrics

In evaluating our model’s performance, we utilized several key metrics. Accuracy, measuring the overall correctness of the model, and the area under the receiver operating characteristics curve (AUROC), which assesses the model’s ability to distinguish between classes, were primary indicators. The optimal classifier threshold was chosen based on the highest precision achieved on the training set. Additionally, we computed sensitivity (recall) to measure the model’s ability to correctly identify positive instances, and specificity to gauge its ability to correctly identify negative instances. The positive predictive value (PPV) and negative predictive value (NPV) were also calculated to provide further insight into the model’s performance.

## IV. Results

We aim to predict evident WPW from lead-I ECG. Here, we report results for the two different methods we explored.

### A. Augmentation and Pretraining

Our first method attempts to learn WPW patterns from examples with only WPW labels against those without any cardiac conditions, i.e. WPW vs Normal. As part of this approach, we explore whether pretraining the model with ImageNet and augmenting in the time domain improves performance. The receiver-operating-curves (ROC) and the area under it (AUC) for the four combinations are shown in Figure 6. These four are the best performing models among the hyperparameter sweeps for the two architectures (ResNet and EfficientNet) described in Section III. Both pretraining and augmentation appear to improve performance on the holdout test set from PTB-XL compared to a randomly initialized model trained without any augmentation. In all cases, however, performances drop when applied on the external Tongji dataset, indicating that augmentation and pretraining do not help models to generalize. Given the large imbalance in the PTB-XL dataset, even the AUC seems to be unable to capture the potential impacts. Hence, we plot the sensitivities and specificities of these models on the two test sets in Figure 7. With pretraining only, though the model’s sensitivity and specificity slightly improves on the internal dataset with a very low presence of WPW examples, both sensitivity and specificity significantly drop on the external set where the number of patients with WPW is much larger. On the other hand, augmentation of the WPW examples seems to improve the performance of the models on both datasets, highlighting the validity of this approach in developing a robust solution.

**Fig. 6:**
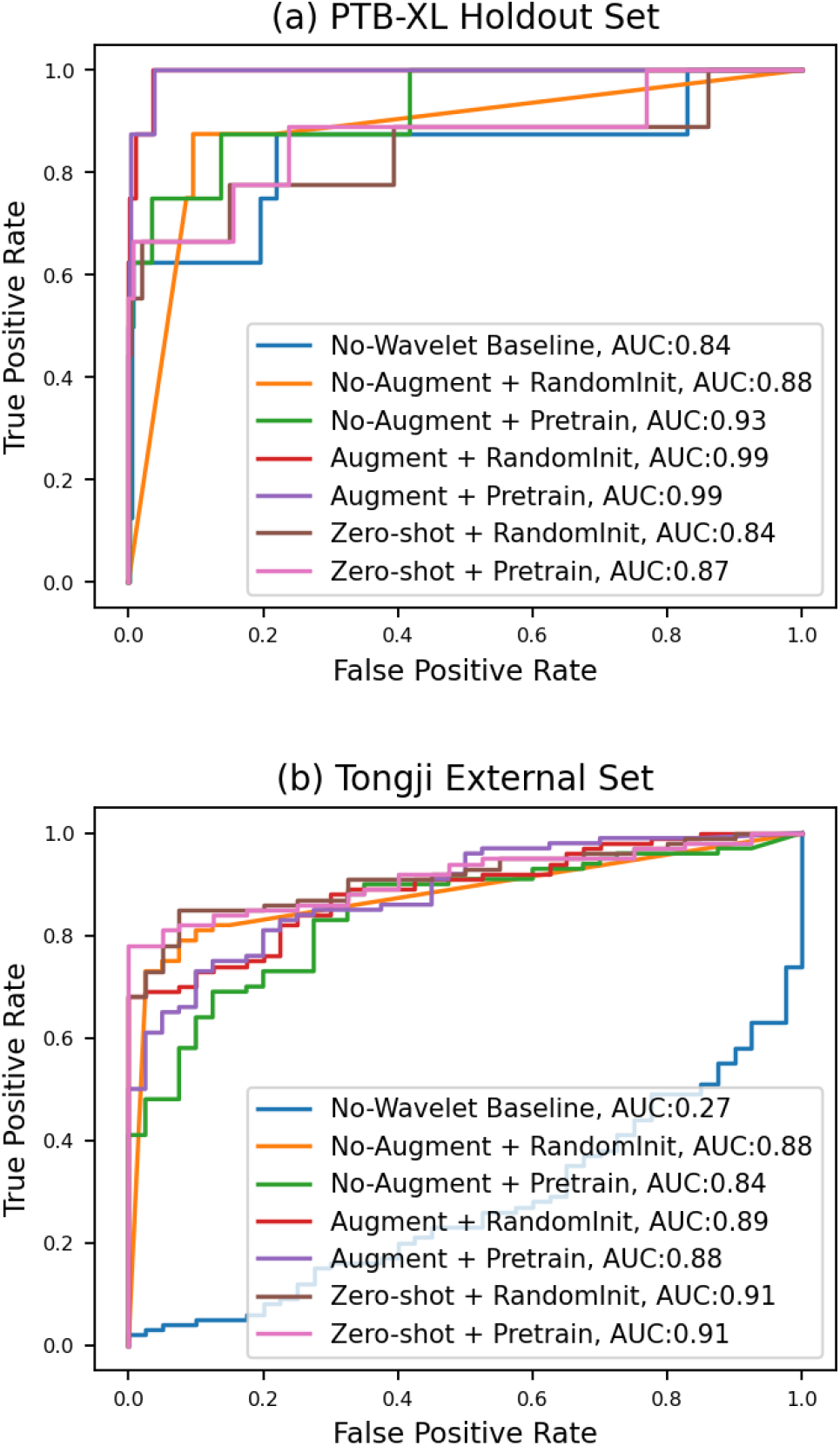
ROC and AUC of different model exploring the effect of various augmentations in identifying WPW against Normal ECG on (a) the internal holdout and (b) the external test-sets.

**Fig. 7:**
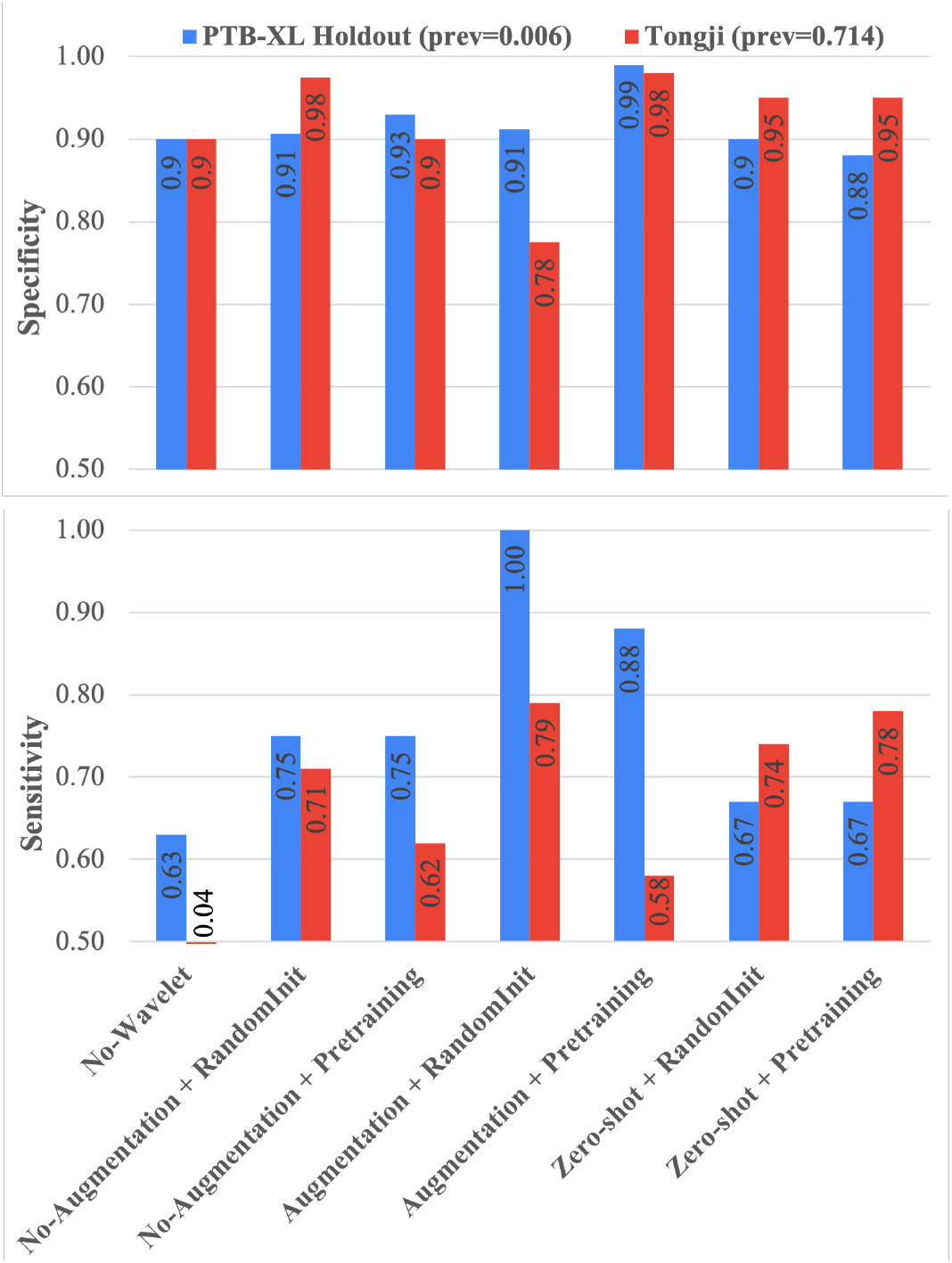
Specificity and sensitivity of different models, without augmentation, with time-domain augmentation, and with the zero-shot transfer learning approach, in identifying WPW against Normal ECG on the internal holdout set and the external Tongji dataset. Model weights are either randomly initialised or imported from pretrained Imagenet-models. All models use wavelet scalogram of lead-I ECG as the input, except the baseline no-wavelet model.

### B. Zero-shot Transfer Learning using CD Superclass

Our second approach attempts to learn CD patterns, instead of WPW only, from examples with CD labels against those without any cardiac conditions, i.e. CD vs Normal ECG. As part of this approach, we explore how such a model trained for a different task performs in identifying WPW samples against normal ECGs. We hypothesize there exists certain ECG pattern similarities amongst WPW and other CD categories, which may work as a natural data augmentation for the WPW class. Hence, we train the model with CD samples, and analyze the model’s predictions to identify the presence of significant separation between the distributions of the logits for the WPW cohort and the normal ECG cohort.

The results for the zero-shot models are presented in Figures 6, 7 and 8. From the AUC plots, we can see only marginal improvement in performance on the holdout PTB-XL test set with pretraining and zero-shot learning (AUC 0.87 vs. baseline AUC 0.84). However, zero-shot performace on the Tongji external set is the highest out of any model (AUC 0.91), indicating that training using the CD superclass provides superior generalization.

**Fig. 8:**
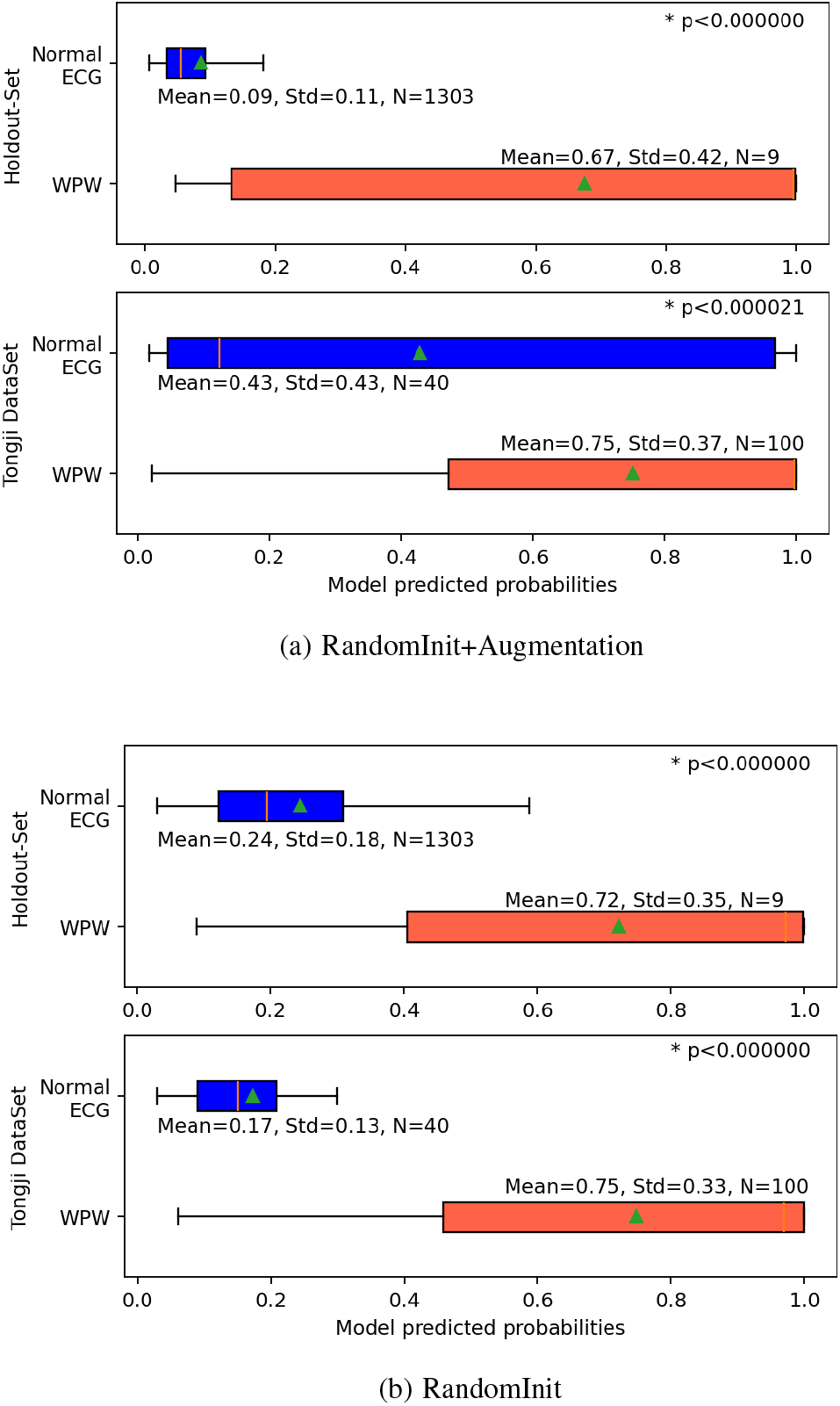
The distributions of the model predictions between WPW and normal ECG cohorts for two randomly initialized (i.e. no pretraining) model configurations: (a) with time-domain augmentation and (b) without augmentation, across the internal and the external testsets. *p-values calculated using Welch’s unequal variance t-test.

From the box-plots of prediction probabilities, we can see that a randomly initialized ResNet model trained to differentiate ECGs with evident CD from normal ECGs assigns significantly lower prediction probabilities to normal ECGs in comparison to WPW cohorts, and thus can be tuned to find a threshold in normal ECG vs those with evident WPW classification tasks.

Also shown in the figure, for a model trained on a relatively balanced dataset, augmentation seems to be less helpful on internal validation, and even harmful for external generalization. The model trained without augmentation is able to assign probabilities for these two cohorts, and the separation between those probabilities are statistically significant. Figure 8b further demonstrates that our transfer learning approach generalizes well, as we notice similar distributions on both the internal holdout test set and the external dataset.

### C. Comparing Wavelet and Time Series Inputs

To quantify the effectiveness of using wavelet-domain scalogram input, we trained both the ResNet18 and EfficientNetb0 using normalized time-series ECG data for both tasks, (1) WPW vs. Normal and (2) CD vs. Normal. In both experiments, the best performing model was using EfficientNetb0 with a batch size of 64 and learning rate 0.0001. The results for this baseline experiment are shown in Figures 6 and 8. We see from the AUC plots that although time-series ECG performs reasonably on the PTB-XL holdout test set (AUC 0.84), it generalizes extremely poorly to the Tongji dataset (AUC 0.27).

Furthermore, we observe that the baseline model has a sharp drop in sensitivity when applied to the Tongji dataset (0.90 to 0.04).

### D. Comparing 12-lead vs lead-I ECG

To evaluate whether the performance drops for lead-I models in comparison to 12-lead solutions, we train a ResNet18 model with 12-lead ECG wavelet input. This 12-lead model achieves 0.91 AUC (vs. 0.88 for the lead-I No-Augment RandomInit baseline, Fig 6) and 0.67 sensitivity (vs 0.75) on the PTB-XL holdout data. On the external Tongji hospital dataset, it achieves 0.90 AUC (vs. 0.88) and 0.69 sensitivity (vs. 0.71). Overall, the 12-lead model seems to slightly outperform and better generalize when compared to the lead-I model.

### E. Why Morse Wavelets

We compare the Generalized Morse wavelet against the Gabor (Morlet) wavelet as well as the short-time Fourier transform (STFT) of lead-I ECG as inputs to our baseline No-Augment RandomInit model. Morse wavelet-based model achieves 0.88 AUC and 0.75 sensitivity on the PTB-XL holdout set and 0.88 AUC with 0.71 sensitivity on the external Tongji dataset. Whereas, Morlet wavelets achieved 0.82 AUC with 0.67 sens on the PTB-XL holdout set and 0.78 AUC with 0.65 sensitivity on the external Tongji dataset. The STFT model achieved 0.85 AUC with 0.67 sensitivity on the PTB-XL holdout set, and 0.86 AUC with 0.55 sensitivity on the external Tongji dataset. Overall, the Generalized Morse wavelet-based model outperforms other wavelet families as well as time-frequency representations.

### F. Technical Contribution

Comparing our work to the results of the PTB-XL baseline experiments conducted by Strodthoff et al is particularly insightful. We report superior performance not only on the PTB-XL holdout test set for the WPW class (0.99 AUC vs. 0.86) but high generalizability to other datasets (Tongji AUC 0.91) using zero-shot transfer learning. Our results seem to confirm Strodthoff’s conclusion that a 1D CNN using time-series input performs reasonably on the internal test-set; however, importantly, we show that these models are very poor at generalizing; in contrast, our wavelet-domain, zero shot models performed very well on external data. This indicates that the CD super-class provided a natural, more generalizable augmentation strategy for classification. Furthermore, our results suggest that wavelet-domain inputs significantly improve the ability of models to generalize to different data distributions, and serve as more robust inputs to deep neural networks than time-series data.

We have also shown that time-domain augmentations improve model performance on holdout test sets, but do not seem to generalize well in our experiments. However, we hypothesize that similar to the approach by Strodthoff, pretraining with PTB-XL followed by fine-tuning on external datasets may somewhat rectify this challenge.

Moreover, our models use only lead-I, whereas previous work on WPW classification has used 12-lead inputs. This is an order of magnitude less data, leading to cheaper training and deployment of deep learning models. Furthermore, this enables application using wearable health devices or in resource-limited settings, in which only lead-I measurements are available. In this regard, our methods could be used as part of ensemble methods for detection of other transient pathologies in lead-I ECGs.

## V. Conclusion and Future Work

In this work, we investigated deep learning-aided automatic diagnosis of WPW from lead-I ECGs. We evaluated several approaches to circumvent model training limitations arising from the rarity of WPW, including time-domain augmentations, ImageNet-pretraining, wavelet analysis, and zero-shot transfer learning using the CD superclass. We evaluated our results both on a within-dataset holdout test set as well as an external dataset, showing both discrimination performance as well as generalizability when compared to previous work.

We assessed of the effectiveness of wavelet analysis with baseline experiments using CNNs on time-series ECG data. The poor performance of these baseline models indicates the efficacy of wavelet analysis in extracting features relevant to WPW identification. We found that time-domain ECG augmentations were useful for holdout set performance, but did not generalize well. More investigation is needed as to identify the inherent explanation of such observations, for example, with pre-training a model using PTB-XL and fine tuning on an external dataset. Finally, we found that zero-shot transfer learning using the CD superclass perform similarly on the PTB-XL holdout test set, but was extremely effective in generalizing to unseen data. By harnessing the electrophysiological similarities within the CD superclass, zero-shot transfer learning offers a promising avenue for developing more flexible and generalizable diagnostic models. We envision that similar approaches can be used for other pathologies with limited data, such as lateral myocardial infarction (LMI, prev. 3 %) and acute MI (prev. 2 %) from lead-I ECG. In PTB-XL, only 201 patients have LMI, in contrast to 2685 inferior MI and 2363 anteroseptal MI patients.

## Data Availability

The study used ONLY openly available human data from PhysioNet: https://physionet.org/content/ptb-xl/1.0.3/.

## VI. Data and Code availability

All code and model weights will be published on GitHub with the final version of the paper. The data used are available on PhysioNet at: https://physionet.org/content/ptb-xl/1.0.3/.

